# Building budgeting capacity of Health Facility Managers to enhance facility financial autonomy: lessons from Nakuru county, Kenya

**DOI:** 10.64898/2026.03.18.26348677

**Authors:** Harrison Ochieng, Fionah Macharia, Joy Mugambi, Peter Nguhiu, Sally Ndungu, Caren Nekesa, Tatiana Ogola, Dennis Amunga, Georges Simiyu, Nelson Kamanda, Wanjiku Chege, Patrick Mwaura, Newton Angwa, Wangari Ng‘ang‘a, Mike Mulongo, Edwine Barasa

## Abstract

**Background:** Health facility financial autonomy enables facilities to retain their revenue and use it to meet facility level needs and priorities to ensure responsiveness, accountability and efficiency. Public facilities need to develop public finance management (PFM) compliant budgets before spending this revenue. However, existing constraints such as lack of competencies and capacities among facility managers in developing budgets and limited political goodwill have influenced the existence of autonomy. This study presents a case study of Nakuru county which implemented an intervention to enhance the capacity of facility managers in developing, implementing and monitoring budgets.

**Methods:** We used a qualitative case study approach, with data collected through participant observations and document analysis. We utilized process evaluation in examining the motivations for the intervention, its implementation, early outcomes and the role of context in these outcomes.

**Results:** The emergence of the intervention was guided by technical, legal and political motivations. The implementation was done in four phases. The first phase targeted the Level four (4) and five (5) facilities who had greater experience with revenue management and already had some level of autonomy, while the second phase built on the lessons learnt and targeted level three (3) and two (2) facilities. The last phase focused on institutionalization and continuous improvement of the standard budgeting process. Early findings showed improvements in budgeting practices in higher level facilities but minimal in level two (2) facilities with some contextual factors such as availability of management staff playing a role.

**Conclusion:** The experience of Nakuru county in building budgeting capacity for facility financial autonomy demonstrates that sustained progress requires a multi-year, adaptive approach that combines training with standardized tools, institutional support, and routine performance monitoring. This journey offers valuable lessons for effective decentralization: tailor support by facility level, embed monitoring and accountability mechanisms, and foster strong leadership and partnerships to sustain gains and enable responsive, autonomous health service delivery.

## BACKGROUND

Facility financial autonomy, a form of fiscal decentralization, allows facilities a level of control and influence to mobilize, allocate and spend financial resources [1,2]. Evidence suggest that this autonomy is an important determinant of facility functioning and performance [3]. For instance, facilities where some level of financial autonomy has been given have shown an increase in finances allocated to procurement of health commodities [4]. Considering these are public funds, there is need for facilities to comply with the different Public Financial Management (PFM) laws which includes the need for budgets that are approved before spending the funds [5]. It is therefore imperative for facility managers to have the necessary knowledge and skills in formulating, executing and evaluating budgets.

In 2010, Kenya enacted a new constitution and transitioned into a devolved system of governance comprising of a central government at the national level and semi-autonomous county governments at the subnational level [6]. In this new arrangement, primary healthcare and county referral facilities are now under the management of the County governments [7]. While these facilities had some level of autonomy pre-devolution, this autonomy was varied post devolution depending on how counties differently interpreted the Public Finance Management Act of 2012 [2]. In some counties, health facilities were required to direct their revenue to the County Revenue Fund (CRF) while in others such as Nakuru County, some level of autonomy was retained without need for additional legislation [3,8].

In 2023, Kenya passed the Facility Improvement Financing (FIF) Act of 2023 that allows facilities to retain their revenue to meet facility needs [9]. Counties such as Nakuru that had retained some level of facility financial autonomy before the passing of this act are now adapting and operationalizing it at the county level. This Act further directs that funds retained by public health facilities are subject to applicable financial laws and regulations [9] including those stated in the Public Finance Management Act (PFM) of 2012 [10] and the Public Procurement and Asset Disposal Act No. 33 of 2015 [11]. Key to this is the need for facility level budgets that are approved before spending to ensure compliance, accountability, transparency and preventing misuse.

Facility managers of Primary Healthcare (PHC) facilities now need to budget for the different funds the facility receives but struggle with existing challenges in their capacity to prepare, report and evaluate these budgets [12,13]. County facilities receive funds from several sources that need to be budgeted for. The county disburses exchequer allocations from the national government to the facilities to help with their running. Apart from these allocations, facilities also receive funds from insurance reimbursements including from the Social Health Authority (SHA), cash payments, grants and donations. These funds need to be budgeted for before spending as required in the PFM Act [10]. However, majority of these managers are often drawn from the clinical staff and have limited financial management training [14]. Additionally, there are other instances where the budgets prepared by the managers are considered “wish lists” as there was limited visibility on ceilings and allocations [15]. Therefore, building the capacity of these managers on budget preparation and management, involvement and transparency in the budgeting process becomes a priority.

Budgeting is a key PFM function and FIF Act requirement that ensures accountability and transparency in the spending of public funds and responsiveness to public needs. Budgeting at the PHC facility level is key in capturing facility-specific needs and health priorities of the population they serve, preventing wastage and enhancing resilience as facilities can promptly respond to shocks [15]. In Kenya, whether and how facilities develop their own budgets varies significantly due to a plethora of factors including differed interpretations of PFM and the FIF Acts, financial and strategic autonomies and the level of trust between county leadership and facilities [5,15]. However, literature suggests that allowing facilities the freedom to develop, manage and report their own budgets can help operationalize and enhance facility financial autonomy reforms [16]. The challenge that remains is that a good number of managers that have a clinical professional background lack in these budgeting and financial management competencies [14].

### County Context

Nakuru county, one of the 47 counties in Kenya has a rapidly evolving population and has made efforts to adapt its health system to respond to the health demands of this population. The county which is located in the rift valley region of Kenya has an estimated population of 2.45 million in 2025 and has undergone rapid urbanization, with nearly 49% of its population living in urban areas following its elevation to city status in 2021 [17]. The county has over 400 licensed health institutions, including major public and private facilities [18,19]. Additionally, the county hosts the Nakuru County teaching and referral hospital whose catchment population extends to the neighboring counties including Baringo, Laikipia, Nyandarua, Kajiado, Narok, Kericho and Bomet. With this population dynamic, efficient use of the county’s financial resources for health is paramount in ensuring that the county effectively meets the healthcare needs of the population it serves.

Uniquely, Nakuru County maintained a level of facility financial autonomy after devolution (2013 going forth), while many other counties centralized facility revenues into the County Revenue Fund (CRF), limiting such autonomy [20]. Nakuru’s approach included early recognition of FIF as a channel for Own Sourced Revenue which is reflected in its County Fiscal Strategy Paper (2017) and in the County Finance Act of 2019 (part V) where some charges were transferred to the FIF, allowing facilities to retain revenue to address operational gaps [21,22] The county is currently domesticating the national FIF Act of 2023 with complementary county regulations enhancing legal accountability and transparent fund management [23]. These reforms are targeted at strengthening and cementing facility financial autonomy.

Regarding budgeting, the county has been transitioning from a consolidated block health department budget to individual facility budgets. In the financial year 2023/2024, the county required Level 4 and 5 facilities to develop their own individual budgets which were to be combined into the health department budget for approval. In the financial years that followed, the county went on to require level 3 and 2 facilities to also develop their own budgets in the 2024/2025 financial year, especially with the coming of Primary Healthcare fund to avoid off-budget spending. Legally, the PFM Act of 2012 and the FIF Act of 2023 require for these facility managers to formulate budgets that are approved by the county before any expenditure [9,10].

The county health department implemented capacity building of these facility managers on budgeting, procurement and reporting to enhance facility financial autonomy. This paper examines the motivations for this capacity building, how it was implemented, the resulting outcomes, the role of context in these outcomes and emerging lessons. This will be useful in providing lessons on subnational government’s engagement in building capacity among health facility managers on the established standardized financial management systems that are necessary for successful facility financial autonomy.

## METHODS

### Study Design

We used a qualitative case study approach, with data collected through participant observations and document analysis. According to Yin, 2017, a case study approach is relevant where the main research questions are “how” or “why”, there is little or no control over behavioral events and the focus of the study is a contemporary phenomenon [24]. Furthermore, the detailed examination of this phenomenon is conducted using the most appropriate nature of the inquiry [25]. We also borrow from process evaluation in the reporting of the findings, highlighting the emergence, implementation, outcomes and the role of context in these outcomes to enhance learning and transferability to other health system contexts [26].

### Study Population

We purposefully selected Nakuru county informed by two reasons. First, this study is part of larger Gates Foundation funded project that is improving PHC performance management in Kenyan counties and Nakuru county is one of our intervention counties. Second, Nakuru county is one of a few counties in Kenya that retained facility financial autonomy post devolution through a Nakuru County Public Finance Management (Hospital Management Services) Regulation, 2014 and was an early adopter of the national FIF Act of 2023 [20]. This financial autonomy of facility managers occasioned the need for budgets and hence the need to build the capacity of managers in preparing these budgets. We used an embedded approach where researchers were co-located in the counties and integrated into the county health system, collaboratively working with the county health department, including supporting the planning and implementation of the budget capacity building intervention. Table 1 presents the characteristics of Nakuru county.

**Table 1:** Key Demographic and Health care information on Nakuru county.

| **Indicator** | **Nakuru County** |
| --- | --- |
| Total Population | 2,162,202 |
| Male | 1,077,272 |
| Female | 1,084,835 |
| Underage 5 | 138,017 |
| Underage 1 | 27,196 |
| **Health Facilities** |  |
| Public | 222 |
| Non-governmental, Faith Based and Private Facilities | 464 |
| **Health Personal-Public Hospitals** |  |
| Total Health Workforce | 3,446 |
| Core Health workforce density per 10,000 population | 16 per 10,000 |
| **Health Financing** |  |
| Total county government health expenditure (FY 2023/2024) | 5,850,962,437 |
| Social Health Insurance Fund (SHIF) coverage in the county (% of county population) | 38% |

Sources: *Kenya Population Census 2019* [27], *Kenya Health Facility Census 2023* [28],
*Nakuru County Health Sector Report 2024* [29].

### Data Collection Procedures

Since the researchers were embedded and actively involved in the planning and implementation of this budgeting capacity building intervention, participant observations with reflexive components were used as a data collection strategy.

Participant observation involves researchers learning about activities of the people or system under study in their natural setting through observing and participating in these activities [30,31]. Data were collected between March 2025 and December 2025 with observations from county health department leaders (FM & JM) that were involved in the implementation of the capacity building, observations from TO & CN that were actively involved in the conceptualization, implementation and evaluation of the capacity building intervention as implementation partners, and an analysis of county level policy and planning documents that had information on the planning and implementation of the intervention. Consent for observations was obtained first from the county health department in the context of implementation of implementation of the primary healthcare performance management program. Second, participants in the capacity building workshops were informed and consented verbally that this was part of action research aimed at improving the effectiveness of this new capacity building approach.

Reflective sessions were conducted between TO, CN, FM, JM and HO in adding depth, learning and credibility to the writing of this manuscript.

### Document reviews

We reviewed content from documents with information on the planning, implementation and evaluation of the budgeting intervention. These documents provided useful information on how the intervention was structured and early findings on the effects of this intervention in improving the ability of the facility managers to develop and implement budgets.

### Data Management and Analysis

Embedded researchers acquired relevant documents relating to the intervention from the county health department and systematically extracted relevant information on how the intervention was implemented (CN & TO). Notes were also recorded during the planning meetings, the actual capacity building meeting and evaluation meetings. In the analysis, we read and re-read the notes and documents to familiarize ourselves with the data. We then developed codes from the data and grouped them to form themes. We triangulated the data from the participant observations and document analysis for refined themes.

### Ethical Considerations

The Scientific and Ethics Review Committee of Africa Medical Research Foundation (AMREF Kenya) gave ethical approval for this work under approval number ESRC p1830/2025. The study has also received approval from the Kenya National Commission for Science, Technology and Innovation (NACOSTI) under license number: NACOSTI/P/25/416429.

## RESULTS

In this section we borrow from process evaluation in discussing what led to the need for the budget capacity building intervention, how the intervention was implemented, early outcomes and the role of context.

### Emergence of the intervention

The intervention was implemented to build the facility manager’s capacity to develop budgets. The department acknowledged that budgets should be made by facility managers since they had information on what the priority facility and population health needs are. It was also a legal requirement in the PFM Act of 2012. Budgeting was also needed to enhance facility financial autonomies that the county was implementing before the national FIF Act of 2023 and was now a legal requirement after the passing of this Act.

While facility managers were required to develop their own individual budgets, there was a significant lack of competencies and capacities. Many facility managers had not been trained in budgeting, procurement or accounting. Majority of these managers were drawn from the clinical staff and had received little to no training in their school curriculum on budgeting and PFM. Therefore, the county aimed to bridge this gap by organizing the sensitization to build the capacity of these managers in budgeting, procurement, and financial reporting procedures, and compliance with PFM guidelines.

There was also political motivation for the implementation of the intervention to implement the governor’s health agenda (2022-2027) in improving the functionality of PHC facilities. The Governor supported interventions that improve the ability of facilities to respond to population health needs. Facilities having budgets and extensive autonomy to implement these budgets ensured accountability, responsiveness and efficiency, aligning with the health agenda.

### Implementation

The capacity-building intervention in Nakuru County was conducted in four technical phases, each addressing facility-specific needs and progressively scaling up support to enhance budgeting capacity and financial autonomy. This multifaceted approach integrated knowledge transfer, practical tools, continuous technical support, and accountability measures to strengthen the capacity of managers to make budgets needed to efficiently and accountably utilize facility autonomy funds.

#### i) Phase 1: Foundations with Level 4 and 5 Facilities

In the financial year 2023/2024, the county department of health insisted on the need for Level 4 and 5 hospitals to have their individual budgets to enhance their responsiveness, accountability and enhance their autonomy. The county recognized that these facility budgets would be best developed internally at the facility level as opposed to the department level because the facility managers better understood the needs, requirements and priorities of the facility. During this financial year, the department brought together facility managers and collaboratively undertook the budgeting process; however, the limited capacity of these facility managers in preparing these budgets was visible. In addition to the facility managers, majority of these facilities had health administrative officers and accountants who could be trained to establish and improve hospital level capacity to prepare, execute, monitor and report budgets.

Based on these realities, the department of health was incentivized to build the capacity of these facility managers, administrative and accounting staff in formulating, implementing and evaluating budgets. In the beginning of the financial year 2024/2025, the department initiated the capacity building intervention by targeting sixteen (16) Level 4 facilities and the Level 5 facility, Nakuru County Teaching and Referral Hospital. The earlier phases of the capacity building started with sensitizing them on why they need to budget and how this should be done. This involved educating them on adherence to the PFM Act of 2012 [10] and the FIF Act of 2023 [9] in the preparation and execution of the budgets, procurement and accounting processes. The facilities were then taken through and adopted a budget template provided by the department of health in line with the county treasury guidelines. This budget template provided for alignment with the county health directoratès programs and therefore made the process of budget consolidation easier and faster. Continuous support was offered to the facility management teams to ensure that subsequent budgets were of the required standard for faster approvals.

#### ii) Phase 2: Expansion to Level 3 Facilities

The capacity building on level 4 and 5 facilities was successful in improving the timelines of budgets, ensuring they reflected facility and county priorities and were compliant with the provided template and PFM requirements. Building on these successes, the capacity-building initiative was expanded to include health centers, engaging Sub-County Health Management Teams (SCHMTs) to deliver localized (per subcounty) training towards the end of the financial year 2024/2025. This was especially important as these facilities now had some form of financial autonomy and were receiving funds from SHA which required budgets that are approved by the county health department before spending. The FIF Act of 2023 had been passed, and the county was in the process of adapting it, which included PFM and FIF compliant budgets at all facility levels. Many Level 3 facilities did not have dedicated accountants and financial management activities were conducted by the facility in-charges who were mainly from a clinical professional background.

As such, the training was modified to include a simplified introduction to PFM and FIF Acts and an opportunity for managers to discuss their concerns and areas they hadn’t understood. Further, the training content was tailored to focus on simplified budget preparation, revenue recognition for smaller transactions, procurement processes, and basic financial reporting.

To ensure sustained support, Level 4 and 5 accountants and health administrators were trained to mentor the Level 3 facility in-charges throughout the budget cycle, thereby strengthening decentralized technical assistance and promoting continuous capacity development.

#### iii) Phase 3: Comprehensive Sensitization for Level 2 Facilities and subcounty health management team

In August 2025 (FY 2025/2026), the county conducted an intensive four-day sensitization targeting 162 Level 2 dispensaries, recognizing their unique challenges such as single-staff operations and limited budgeting experience. Like level 3, these facilities were also now receiving funds from SHA which also needed to be budgeted for. The managers were sensitized on the need to adequately and transparently plan, spend and report public funds as enshrined in the PFM Act of 2012. The training emphasized standardized budget templates, practical exercises, peer learning, and detailed stepwise guidance on budget preparation. Objectives included sensitizing on county budgeting processes and legal requirements (PFM Act 2024 amendment and FIF Act 2023), budget preparation, financial reporting and procurement planning to promote transparency and accountability in fund use. The Subcounty team leads and the health management teams were also included in the training, recognizing their important role in supporting Level 2 managers in planning and budgeting.

#### iv) Phase 4: Institutional Support and Continuous Improvement

Beyond training, the department established institutional mechanisms to ensure sustainability. Standardized budgeting templates continued to be used at the facility level to ensure compliance, monthly performance reviews were introduced to monitor revenue tracking, timely issuance of Authority to Incur Expenditure (AIE), and submission of Social Health Insurance (SHA) claims. A county budget core team was designated to review performance data and provide targeted follow-up support, while structured feedback loops were created to systematically assess impact and inform subsequent budgeting cycles. This continuous monitoring was important in holding the facility managers accountable and identifying issues that were emerging or had been missed out.

### Monitoring outcomes of the capacity building

#### Improvements in budgeting processes and capacity

Monthly performance reviews conducted by the County Department of Health in the financial year 2024/25 and early 2025/2026 highlighted varying outcomes across facility levels to track budgeting practices. Level 4 and 5 facilities demonstrated improvements in their budgeting practices which included the timeliness and accuracy of their budgets, alignment with PFM requirements and capturing of facility and county priorities. These higher-level facilities had accounting and administrative staff and medical superintendents who were trained in budgeting and hence the budgeting roles were distributed across them. This possibly explains the better uptake and application of the knowledge and skills gained from the capacity building intervention.

Level 3 facilities also showed improvements in the timeliness of their budgets; budgets were submitted on time and were in adherence to the standardized templates that had been provided which also fastened the approval process.

In contrast, the change in Level 2 facilities (dispensaries) was slower compared to the other levels due to continued dependence on sub-county managers for budgeting, limited staffing, and competing clinical and administrative responsibilities. A summary of these findings is shown in table 2 below;

**Table 2;.**
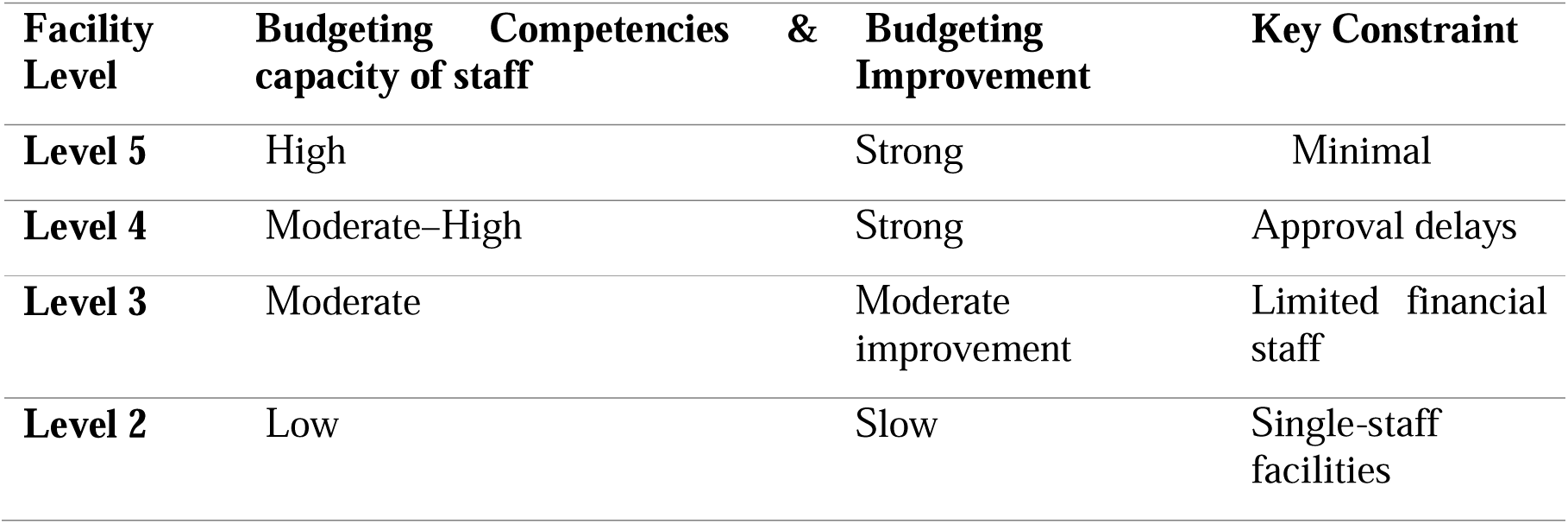
Improvements in budgeting practices across facility levels.

Overall, the findings indicate that capacity building yields the greatest impact where enabling systems and structures are already in place. Structural barriers at lower-level facilities constrain results, though standardized tools have accelerated adoption and ongoing monitoring continues to promote incremental improvement. Historical institutional contexts also played a key role in shaping performance outcomes.

#### Addressing facility pending bills

Considering there were improvements in the knowledge and understanding of facility managers and accounting staff in the budgeting process, there was an opportunity for an informed discussion on how to address the challenge of pending bills. Facilities were required to allocate 30% of their budget to development. However, the department and the managers of Level 4 and 5 facilities agreed that to effectively tackle the challenge of pending bills, 15% of the FIF budget would be allocated to settling pending bills and the other 15% to development in the financial year 2024/2025. The same resolution was adopted by Level 3 facility managers in the financial year 2025/2026. The department reported that there was now progress by facilities in meeting pending bills.

### Impact of context on the outcomes

The existing political support for autonomy reforms was an enabler to the implementation and outcomes of the intervention. Nakuru county is one of the pioneering counties for facility autonomy post devolution (after 2013). The county has also been an early adopted of the national Facility Improvement Financing (FIF) law and is in the process of developing county specific FIF law and regulations to better domesticate this act. The current county government leadership intended to enhance this facility autonomy to enhance service delivery at the facility level for improvements in health outcomes. This political support further encouraged the county health department’s technical duty to enhance facility financial autonomy by building the capacity of facilities to develop, manage and report their own budgets.

The involvement of implementing partners provided technical support and catalyzing funds for the implementation of the intervention. Technical partners including Hecta were actively involved in the planning, implementation and evaluation of the capacity building. These partners who had experience in budgeting, strategic purchasing and PFM provided technical guidance on what to include in the capacity building, how to effectively deliver the training and means to monitor and evaluate the outcomes. Additionally, they also offered catalytic funding to meet some of the initial costs for implementing the intervention while county provided the remainder of the funds. This funding approach was sustainable and “set the wheel in motion” in the long term.

The shortage of staff at the level 2 facilities slowed down progress of the intended outcomes. A good number of level 2 health facilities were managed by one clinical staff who had to “juggle” between clinical and administrative duties. In cases where the workload was high, these managers took a lot of time to attend to patients, limiting the time they had to prepare the budgets. As such, what was observed is that budgeting at this level was still not timely and to the quality required. However, the county health leadership perspective was that this was not a capacity challenge but rather the fact that the managers had limited time to sit down and develop budgets to the quality required.

### Emerging Lessons

Nakuru’s experience underscores the value of initiating budgeting capacity development early and sustaining it through a multi-year, adaptive approach. Standardizing tools such as budgeting templates created and shared prior to scaling training proved critical for consistency and effectiveness. Support should be context specific. Higher-level facilities benefit from periodic reinforcement, while lower-level, single-staff dispensaries require continuous, hands-on assistance and simplified financial processes. Embedding routine monthly monitoring strengthens accountability and promotes continuous improvement.

Addressing structural barriers, such as staffing shortages, demands targeted strategies that extend beyond training interventions. Sustained capacity building depends on strong county leadership, strategic partnerships, and leveraging existing informal practices.

Key success factors include collaborative development of standardized tools, integration of performance monitoring into regular management processes, and a commitment to adapt approaches based on evidence. Common pitfalls to avoid include assuming training alone can resolve systemic challenges, applying uniform strategies across diverse facility contexts, implementing one-off interventions without follow-up and conducting monitoring without responsive action.

## DISCUSSION

The County department of health working with partners implemented a budgeting capacity building intervention to enable managers to effectively prepare and execute budgets. This was in response to the identified knowledge and skills gap among facility managers who were primarily clinicians. The capacity building initiatives were implemented in phases, starting from higher level facilities and cascading to lower-level facilities. Monthly reviews showed these higher-level facilities demonstrated improved budgeting capacities, but the progress was slower in lower-level facilities due to staff shortages and competing administrative responsibilities. Though mixed, these improvements will enable facility managers to effectively manage their budgets transparently, efficiently, and in line with the PFM and FIF Acts.

Literature across LMICs shows facility managers mostly drawn from the clinical area often have limited capacity in preparing and managing budgets, a gap this capacity building intervention was aiming to address. Majority of the facility managers in the county were doctors, nurses and other clinical cadres who had little to no training in budgeting and financial management. A study conducted by Wamalwa et al., 2023 in Bungoma county, Kenya found that 58% of health managers including facility managers had not received any training on the annual planning process which informs budgets and 81% had not been trained on Program based budgeting (PBB) [12]. Another study conducted in Zambia by Foster et al., 2017 also found that nurses in managerial roles had received little training on preparing facility budgets during in their college curriculum [13]. Therefore, addressing these knowledge and skill gaps through interventions such as this by Nakuru county becomes integral to the transparent and efficient use of resources and effectively enabling and implementing facility financial autonomy.

The county is showing good political support for facility autonomy reforms by implementing this budgeting capacity building activity to strengthen the transparent, accountable and efficient use of FIF funds, contrary to what has been documented in Kenya and the region. Evidence from Kenya has shown that in some counties, county leaders have a political interest to consolidate their power and retaining control over these facility generated revenue is a means to achieve this [3]. This is similar to what was seen in Malawi where political actors were against hospital autonomy as they perceived it as a loss of power and control and “potential loss of patronage resources” [32]. This little to no political support for autonomy reforms has therefore been a barrier to facility financial autonomy despite its documented efficiency and accountability gains [1,3,32]. Nakurùs case provides an example where the political interests are aligned towards effective service provision as captured in the “Governor’s Health Agenda” with the intention it translates to political support from the public. This appreciates the important role of political support in accelerating facility management capacity and autonomy. Health leaders and other technical partners should also consider seeking this support and goodwill when planning and implementing facility autonomy reforms.

The capacity building intervention for lower-level facility managers was implemented in recognition of their local context and constraints rather than applying blanket principles across all facility levels. According to the 2023 Kenya health facility census report, only 1 % of level 2 facilities and 2% of level 3 facilities had all the requisite health cadres [28]. From this census, 80.4% of level 2 facilities in Nakuru county had less than 3 nurses. Our current work in these counties has also shown that Level 2 facilities are the most affected by staff shortages with some facilities only having one nurse who carries out clinical and administrative duties. Recognizing this challenge, the county health department tailored the training for level 2 facility managers to one with more simplified content compared to that used for the higher-level facilities, reduced to only what was necessary in regard to PFM, planning and budgeting, including practical exercises, peer learning, and detailed stepwise guidance on budget preparation. This approach of shortening to only what is necessary, using practical exercises and mentoring are best practices in ensuring success of management capacity building at the facility level [33].

Under the Primary Care Network (PCN) guidelines, subcounty health management team is expected to provide leadership and management support to health centers and dispensaries within their hub [34]; literature has also shown that managers from these facilities have sought guidance from the subcounty health management team on matters relating to facility financial reforms [35]. As such, the inclusion of the subcounty health management team in the training was integral in establishing a support system for these lower-level facility managers. These approaches, alive to the local realities, ensured that the capacity building was useful and practical for these managers

Higher level facility and subcounty health management teams can mentor and offer technical support to lower-level facility managers to build their capacities and ability to carry out the budgeting process on their own. Lower level facilities have few staff and hence run the challenge of not effectively implementing these budgeting and other facility autonomy reforms as discussed by Witter et al., 2025 [16]. However, this case provides evidence on how existing administrative arrangements from higher level facilities and subcounty can be leveraged to provide the needed support to these facilities as they “settle into” these reforms. Nakuru county has primary care networks (PCNs) which are made of spokes (level 2 and 3 facilities) and hubs (level 4s and 5 facility). The department utilized this structure in pairing level 2 and level 3 facility managers with managers from the higher-level facility and the subcounty health management teams. Supportive supervision and mentorship from higher (district or county) level leadership to facility managers has been shown to be effective in improving staff attitude, motivation and facility performance [14,36]. As such, this case study provides an example of how such an approach can be utilized in building the financial management capacities of lower level facility managers.

Drawn from our findings and discussion relating it to literature, we make the following recommendations. One, Counties and partners should institutionalize ongoing capacity building tailored to facility levels, combining standardized budgeting tools and context-specific support to address unique challenges, particularly for resource-constrained dispensaries to strengthen financial autonomy and improve service delivery. Secondly, Counties implementing the FIF Act of 2023 should integrate financial performance monitoring and accountability into routine processes, such as regular performance reviews. This approach enables early identification of gaps, supports timely budgeting, resource allocation, and AIE approvals, and fosters continuous improvement in financial management.

### Study Limitations

Being a qualitative case study, we acknowledge that our study is limited in not being able to accurately show causality between the improved budget processes and service delivery changes. Further studies can employ mixed methods to assess this.

## Conclusion

Nakuru County’s experience in building budgeting capacity for facility financial autonomy demonstrates that sustained progress requires a multi-year, adaptive approach that combines training with standardized tools, institutional support, and routine performance monitoring. Success was greatest where facilities had established financial systems and dedicated staff, while lower-level dispensaries faced persistent challenges due to limited staffing and competing responsibilities, highlighting that capacity building must be paired with strategies addressing structural barriers. The county’s journey offers valuable lessons for effective decentralization: tailor support by facility level, embed monitoring and accountability mechanisms, and foster strong leadership and partnerships to sustain gains and enable responsive, autonomous health service delivery.

## Acknowledgements

We would like to acknowledge the County Department of Health for their relentless commitment to improving budgeting capacity at the facility level. We would also like to acknowledge other partners who are participating in making this work a success; the Financing Alliance for Health, ThinkWell and AMREF

## Ethics Statement

Ethical approval was sought from AMREF Scientific and Ethics Review Committee, and ethical approval was obtained; approval number ESRC p1830/2025. The study has also received approval from the National Commission for Science, Technology and Innovation (NACOSTI), license number: NACOSTI/P/25/416429.

## Data Availability Statement

All data that are relevant for peer review and learning are available in the manuscript.

